# Identification of cellular senescence-related gene *IFNG* as a potential biomarker in acute rejection after kidney transplantation via weighted gene co-expression network analysis and multiple machine learning

**DOI:** 10.1101/2025.06.19.25329910

**Authors:** Changhai Xu, Xueying Wang, Haibo Wu, Wei Li, Fei Lin, Na Lin, Shiyin Shen, Shubin Pan, Tong Chen, Donghui Zhang, Long He, Yan Cui

## Abstract

**Background:** Kidney transplantation is the best option for the treatment of end-stage kidney disease (ESKD). Acute rejection (AR) episodes are a major determinant of renal allograft survival. Cellular senescence in the pathogenesis of allograft failure. Herein, we aimed to explore hub cellular senescence-related gene in AR after kidney transplantation.

**Methods:** The data used in this study was obtained from the Gene Expression Omnibus database. The hub cellular senescence-related gene was identified using WGCNA and three machine learning algorithms. The function information was analyzed using the GO and KEGG enrichment analysis. The correlation between hub gene and immune cells was calculated using ssGSEA algorithm and Pearson’s correlation analysis.

**Results:** A total of 31 cellular senescence-related genes were differentially expressed in the AR and stable groups. Among which, 19 genes were correlated with onset of AR after kidney transplantation. After utilizing the three machine learning algorithms, *IFNG* was identified as the hub cellular senescence-related gene. *IFNG* was highly expressed in AR samples, and it could better distinguish between stable individuals and AR patients after kidney transplantation. Moreover, the expression of *IFNG* was closely correlated with immune cell infiltration and function. *IFNG* expression was associated with multiple drugs. Finally, we found that *IFNG* was high expressed in kidney tissues of AR in allogeneic kidney transplant mice

**Conclusions:** Our study revealed that cellular senescence-related gene *IFNG* might be a potential biomarker AR after kidney transplantation.

## 1 Introduction

Kidney transplantation is well-established as the best treatment for end-stage kidney disease (ESKD) [1]. Due to progress in surgical methods, the extensive use of tissue compatibility assessments, the arrival of perioperative antibody induction therapies, and the creation of new and effective immunosuppressive agents, the average survival rate of renal transplants has greatly enhanced [2–4]. However, rejection, particularly acute rejection (AR), remains a major challenge that impacts the long-term survival of transplanted kidneys [1, 3, 4]. AR is the most prevalent form of rejection following kidney transplantation and primarily occurs in the early stages post-transplant. It affects 15-20% of kidney transplant recipients and significantly influences long-term graft survival [5–7]. Consequently, AR should be a key consideration in the differential diagnosis of unexplained graft dysfunction in transplant recipients.

Multiple mechanisms have been proposed to be involved in AR after kidney transplantation pathophysiology, such as T cell-mediated rejection (TCMR) [8], antibody-mediated rejection (ABMR) and ischemia-reperfusion injury [9]. The pathogenesis of TCMR involves the activation and proliferation of T cells, which directly contribute to the destruction of transplanted kidney tissue [10]. These activated T cells can release cytokines that trigger inflammatory reactions, ultimately resulting in damage to the transplanted kidney [10]. AMR primarily results from the interaction between recipient antibodies and donor organ antigens [11]. Antibodies attach to surface antigens on transplanted kidney cells, activating the complement system, which in turn leads to cell lysis [12]. Alternatively, these antibodies may bind to surface antigens on the vascular endothelial cells of the transplanted kidney, causing damage to these cells and resulting in tissue ischemia and necrosis [12].

Cellular senescence is a state in which cells cease to proliferate, leading to inflammation, impaired tissue repair, irreversible tissue damage, and organ dysfunction [13]. Recent studies have implicated cellular senescence in the pathogenesis of allograft failure [14]. Renal cell senescence plays a causative role in the development of interstitial fibrosis, tubular atrophy, and renal allograft degeneration by preventing recovery after injury [15]. Furthermore, senescence marker, p16(INK4a) is highly expressed in deteriorating renal transplants and diseased native kidney [16]. However, the relationship between cellular senescence and AR after renal transplantation is complex and currently unclear. Therefore, it is of great significance to elucidate the role and mechanism of cellular senescence in the development and progression of AR after kidney transplantation.

To systematically evaluate the correlation between cellular senescence and the pathogenesis of AR in kidney transplantation, we identified *IFNG* as a novel cellular senescence-related gene (CSRs) in AR after kidney transplantation via weighted gene co-expression network analysis (WGCNA) alongside multiple machine learning algorithms. We also assessed the potential mechanisms through which IFNG may influence AR after kidney transplantation. Subsequently, we analyzed the relationship between *IFNG* expression and immune cell infiltration in patients undergoing acute renal transplant rejection. This study provides new scientific basis for the prevention and treatment of kidney transplant rejection by revealing the relationship between cellular senescence and AR after kidney transplantation.

## 2. Materials and Methods

### 2.1. Subjects

Six databases on post-kidney transplant AR and stable graft (STA) samples were found in the Gene Expression Omnibus (GEO) (http://www.ncbi.nlm.nih.gov/), including GSE25902 (AR =24, STA=96), GSE14328 (AR=18, STA=18), GSE50058 (AR=43, STA=58), GSE129166 (AR=18, STA=18), GSE53605 (AR=58, STA=154), and GSE36059 (AR=100, STA=281).

A total of 949 human CSRs [17, 18] were derived from the CellAge [19] (**Table S1**) (https://genomics.senescence.info/cells/).

### 2.2. Differentially expressed gene analysis

The R package limma (version 3.5.2) [20] was employed to identify differentially expressed genes (DEGs) between two groups using the thresholds of |log2FC| > 0.5 and p.adjust < 0.05.

### 2.3. Functional enrichment analysis

The R package Clusterprofiler (version 4.8.3) [21] was used to perform Gene Ontology (GO) and Kyoto Encyclopedia of Genes and Genomes (KEGG). The “DOSE” [22] was performed Disease Ontology (DO) enrichment analysis. A p-value <0.05 was considered statistically significant.

### 2.4. Gene set variation analysis (GSVA)

The R package GSVA [23] was used to perform GSVA across groups and calculate enrichment scores. Subsequently, differential analysis between groups was conducted using the R package limma ( version 3.56.2) [20], and pathways with p.adjust < 0.05 were selected as significantly enriched.

### 2.5. WGCNA

The R package WGCNA [24] was utilized for WGCNA analysis based on gene expression values. Genes were filtered using variance analysis to retain the top 75% for WGCNA. Pearson correlation coefficients between genes were computed, and an appropriate soft threshold β was selected to ensure the network approximates a scale-free network. A one-step approach was employed to construct the gene network, converting the adjacency matrix to a Topological Overlap Matrix (TOM). Hierarchical clustering was performed to generate a dendrogram. Gene significance and module significance were calculated to assess the association between genes and clinical information, and significant correlations between modules and traits were analyzed to gain gene modules. The intersection of Hub genes and DECSRs in the module was taken to obtain hub genes.

### 2.6. Identification of hub genes with highly correlated AR features

LASSO regression analysis was performed using the “glmnet” package in R [25]. Decision tree construction and feature selection were conducted using the random forest method with the “randomForest” package in R. Variable selection was further refined through recursive feature elimination using the “caret” package in R.

### 2.7. Immune cell infiltration analysis

The “GSVA” package [23] was used to calculate enrichment scores for 28 immune cell types [26] and 13 immune functions [27].

### 2.8. Animals

This study utilized a total of 12 C57 mice aged 6 to 8 weeks, all of which were housed in a clean and ventilated animal room to acclimate to their environment. Prior to the experiment, all mice underwent a fasting period of 12 hours with free access to water. Thirty minutes before the procedure, heparin sodium was administered via intramuscular injection to prevent thrombosis. Euthanasia was performed by the cervical dislocation method, and isoflurane was used as the anesthetic during surgery, with an initial concentration of 6% (flat rate: 1L/min) for induction and a maintenance concentration of 2% (flat rate: 0.8-1L/min). All animal-related experiments were conducted in accordance with the National Guidelines for Ethical Review of Laboratory Animal Welfare. Ethical approval for all mouse experiments was obtained from the Laboratory Animal Ethics Committee of Beijing MDKN Biotech Company (MDKN-2023-054).

### 2.9. Mouse kidney transplantation

Donor Surgery: Following the successful administration of anesthesia to the mouse, a midline abdominal incision was made to fully expose the left kidney of the donor, including the vascular, ureteral, and bladder regions. The left gonadal vein was ligated and severed, while the adrenal arteries and veins were electrocoagulated and subsequently divided. The abdominal aorta and inferior vena cava were dissected 2 mm below the left renal vessels, and the inferior vena cava was transected approximately 5 mm below the left renal vessels. Careful cauterization and dissection were executed on the gonadal vessels, which were formed by the left lumbar vein, inferior vascular branches, and renal pedicle. A volume of 0.5 to 1 ml of cold heparinized saline was slowly perfused until the left kidney exhibited a pale appearance. The abdominal aorta and vena cava were transected above the ligature, and the ureter was cut at the mid-segment of the left renal ureter. The kidney was completely liberated from surrounding tissues, and the left kidney, ureter, and bladder were removed en bloc and placed in ice-cold saline at 4°C for preservation.

Receptor surgery: After anesthesia, the mice were fixed on the surgical plate, and the abdominal surgical area was sterilized and prepared for skin. A median subxiphoid to suprapubic incision was made to expose the right kidney region, and the right kidney was resected by ligating and dissecting the right ureter and renal hilum. The left renal inferior vena cava and abdominal aorta were exposed, the lumbar artery was ligated, and the IVC and AO were occluded with visible microvascular clips at a distance of 4-5 mm. The transplanted kidney was placed on the right side, the renal artery was anastomosed end to end with 11-0 nylon suture and the renal vein was anastomosed end to end with 10-0 nylon suture under a 15X surgical microscope, and urinary tract reconstruction was performed after opening the blood vessels. The donor bladder was trimmed under a 10X surgical microscope, the wall of the bladder without blood supply was cut obliquely and removed, and the recipient bladder was trimmed to open the opening, and the donor bladder flap was anastomosed with a continuous full-layer anastomosis of the donor bladder flap and the recipient bladder with 10-0 nylon thread, and the bladder was filled with water to check that there was no leakage. After fixation of the transplanted kidney and repositioning of the bowel, 1 ml of warm saline was left in the peritoneal cavity, and the peritoneal cavity was closed in two layers with 0 silk suture.

After surgery, the mice were placed in warm, clean animal cages and monitored for vital signs and healing of surgical incisions. Antibiotics were administered to prevent infection, and the weight and urine output of the mice were monitored regularly to observe changes in vital signs such as respiration, heart rate, and blood pressure. The surgical incision was monitored for infection, bleeding, and poor healing.

### 2.10. RT-qPCR

RNA extraction from the tissues was performed using TRIzol reagent (Invitrogen, USA). Reverse transcriptase (Vazyme Biotech Co., Ltd.) was utilized to convert the RNA into complementary DNA (cDNA), which was then analyzed through an RT–qPCR assay with ChamQ Universal SYBR qPCR Master Mix (Vazyme Biotech Co., Ltd.). The quantified results were normalized against the endogenous levels of GAPDH expression. The primer sequences used in the RT–qPCR assay can be found in Table 1.

**Table 1.**
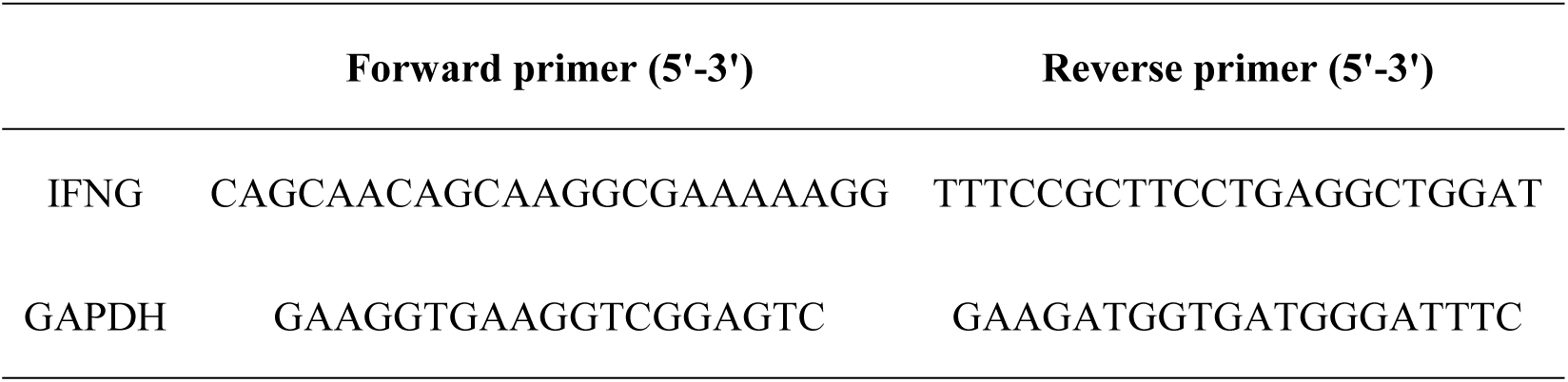
The sequences of the primers used for RT–qPCR

### 2.11. Western blot assay

Proteins extracted from kidney tissues were separated via SDS–PAGE and then transferred to PVDF membranes (ab133411, Abcam). These membranes were incubated with the primary antibody at 4 °C for 12 hours, followed by a one-hour incubation with secondary antibodies. The primary antibodies utilized in this research included Anti-IFNG (ab122397, Abcam) and Anti-GAPDH (ab8245, Abcam). The secondary antibody employed was Goat Anti-Rabbit IgG H&L (ab150077, Abcam). Visualization was performed using the SuperPico ECL Chemiluminescence Kit (E422-01/02, Vazyme) on an Odyssey Infrared Imaging System (LI-COR Biosciences, USA). The optical densities of the membrane blots were assessed using ImageJ software. The primary images of the blots are showed in Figure S1.

### 2.12. Histological evaluation

Kidneys were fixed in 10% neutral-buffered formalin and subsequently embedded in paraffin. The paraffin sections, each 3 μm thick, were stained with hematoxylin and eosin (H&E). Following staining, the slides were dehydrated using a series of ethanol gradient washes and clarified with xylene washes. The sections were then mounted with neutral resin and observed under a microscope, where they were photographed.

### 2.13. Statistical analysis

The Wilcoxon rank-sum test was used to compare gene expression differences and immune cell infiltration differences between groups. Pearson correlation analysis was conducted using R. Statistical significance was defined as a p-value of less than 0.05. All statistical computations were performed with R software (version 4.3.3).

## 3. Results

### 3.1 Identification of differentially expressed CSRs (DECSRs) between AR and STA samples

To analyzed the DECSRs between AR and STA samples, we first merged GSE55235, GSE55457, and GSE12021 databases after removing the batch effects via the “sva” packages in R packages (Fig. 1A, 1B). The merged dataset contains 172 STA and 85 AR samples and was named GEO-meta dataset. In the GEO-meta dataset, we found that 31 DECSRs were significantly differentially expressed between AR and STA samples (Fig. 1C, Table S2), and the heatmap of gene expression is shown in Fig. 1D.

**Fig. 1.**
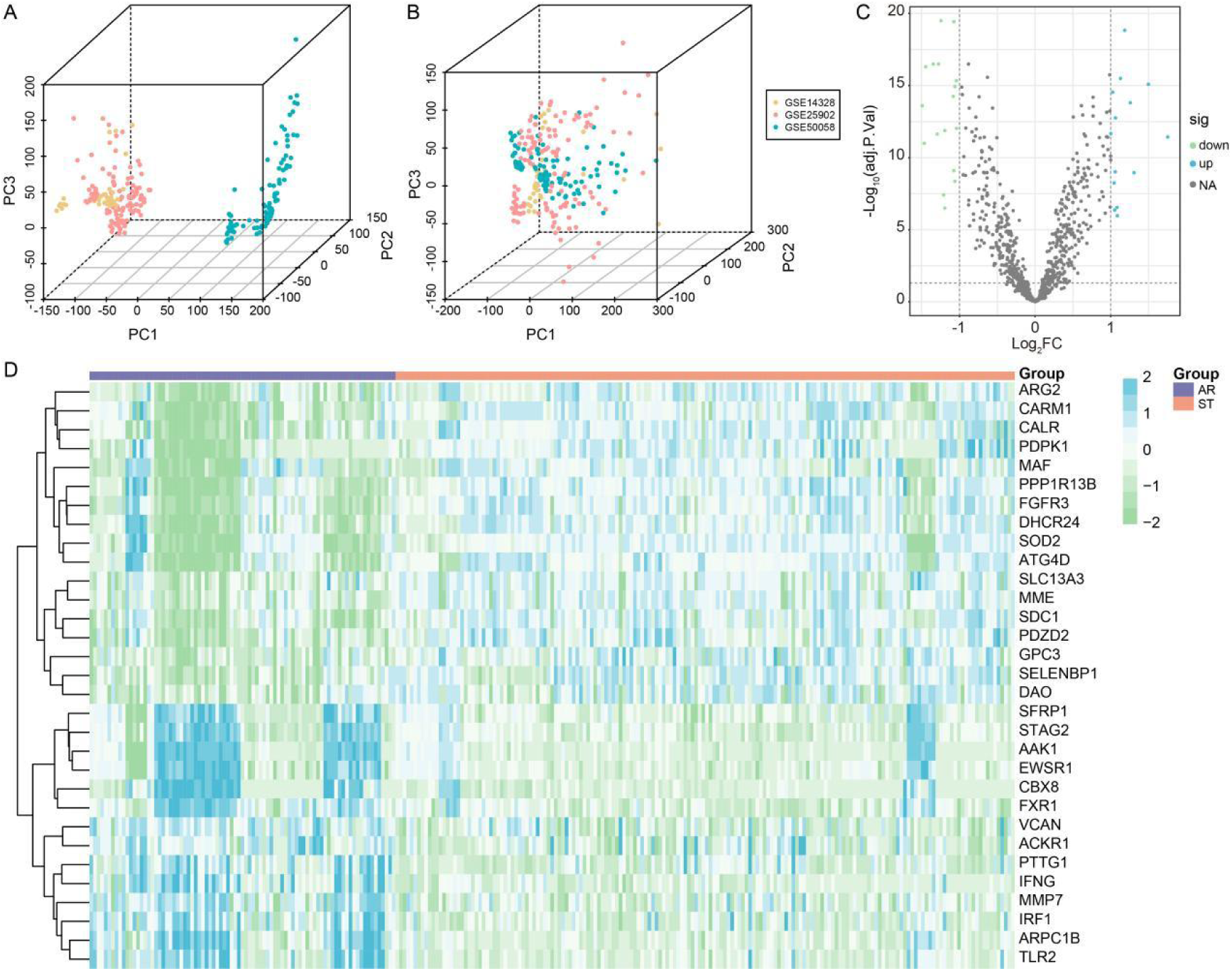
Ldentification of differentially expressed cellular senescence-related genes in AR samples. **(A).** Principal component analysis before samples elimination of batch effects. **(B).** Principal component analysis after samples elimination of batch effects. **(C).** Volcano plots of gene expression differences between the GEO-meta dataset before and after de-batching effects. **(D).** Heatmap of gene expression between before and after de-batch effects in the GEO-meta dataset.

To better understand the potential mechanisms of 31 DECSRs in AR, we performed GO, KEGG, and DO enrichment analysis on 31 DECSRs. GO enrichment analysis revealed 90 significantly enriched BP terms, and 4 significantly enriched CC terms (Fig. 2A). Furthermore, KEGG pathway analysis revealed that these DECSRs were enriched in the malaria and inflammatory bowel disease interacted closely (Fig. 2B). The Fig. 2C depicted the top ten DO enrichment pathways, such as bone marrow cancer, organ system benign neoplasm, and myeloid neoplasm. Table S3 showed detailed results for GO, KEGG, and DO enrichment of DECSRs.

**Fig. 2.**
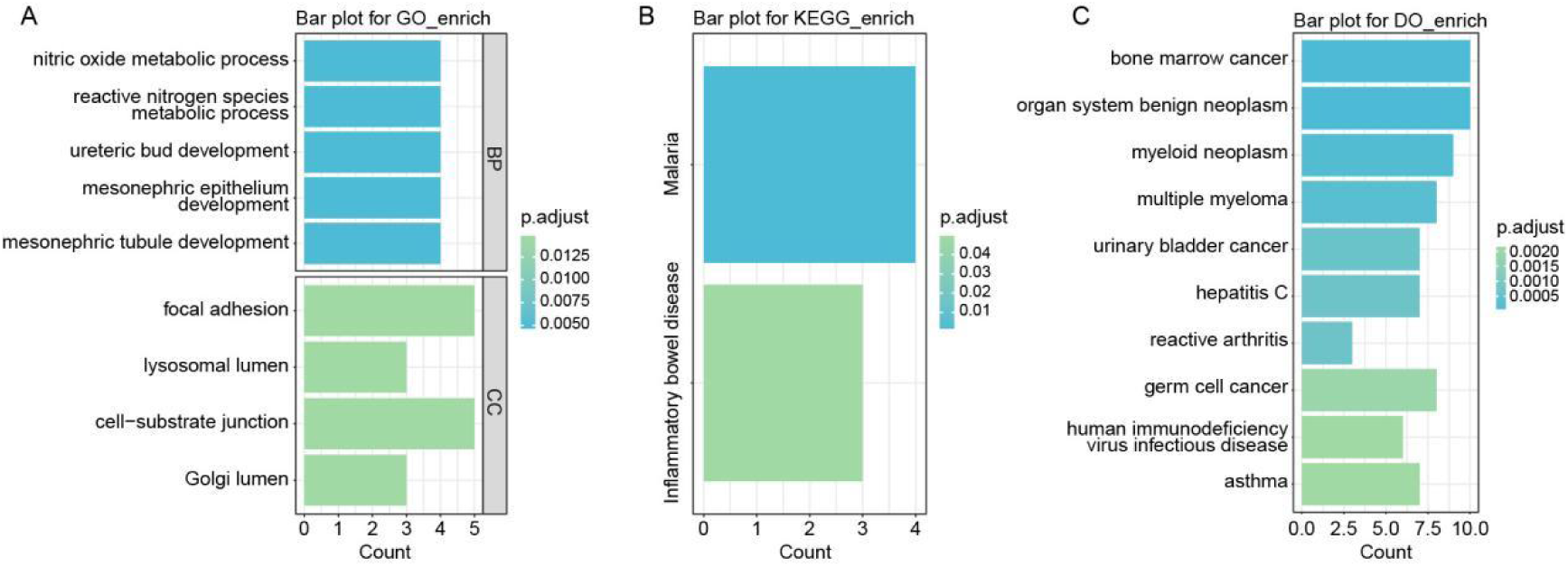
Functional enrichment analysis of DECSRs. **(A).** The most significantly enriched 5 BPs, 5 MFs, 5CCs in GO enrichment analysis. **(B).** The most significantly enriched KEGG pathway. **(C).** The top 10 most significantly enriched DO pathways.

### 3.2 Identification of AR onset related genes

We performed WGCNA using the GEO-meta dataset, and filtered the top 75% of genes for WGCNA analysis. The value 12 was selected as the optimal soft threshold to construct the gene network (Fig. 3A). A total of 11 gene modules were obtained (Fig. 3B). Based on the AR and STA information in the cohort, they were made as trait data for WGCNA, and the correlation between each gene module and the two types of samples was calculated (Fig. 3C, 3D). We selected four the most related modules with *p*-values less than 0.05 (MEblue, MEgreenyellow, MEblack, MEyellow) and obtained a total of 3656 genes as candidate genes (Table S4). Cross-analysis of the above DECSRs with 3656 genes yielded a total of 19 AR-DECSRs (Fig. 3E, Table S5).

**Fig.3.**
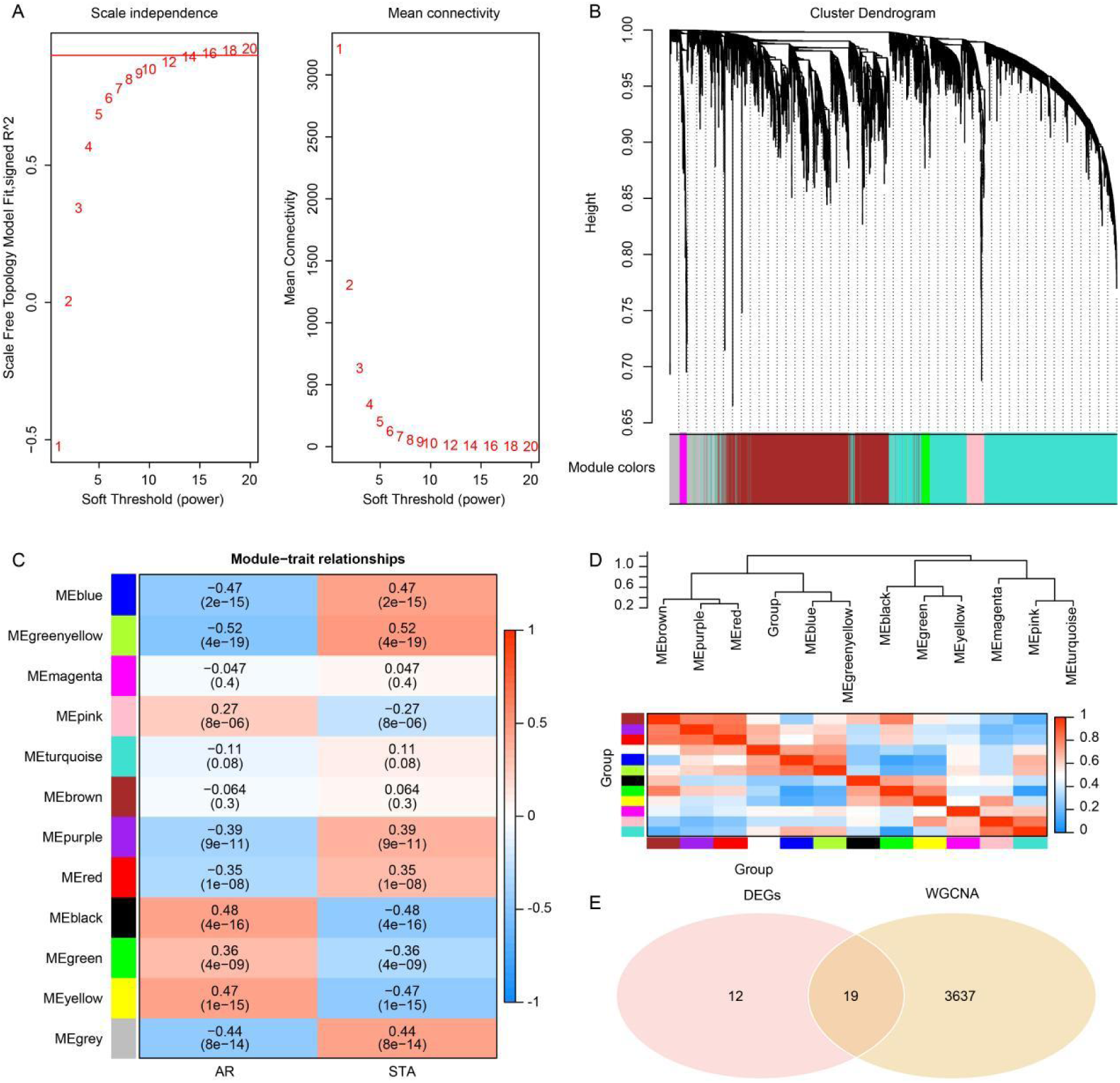
WGCNA. **(A).** The soft threshold value of 18 was determined as the best soft threshold. **(B).** The variance is in the top 75% of the gene cluster dendrogram, with each branch of the graph representing a gene and each color below representing a co-expression module. **(C).** Heat map of module-trait relationships, where each color represents a co-expression module and the values represent module-trait correlation coefficients and p-values. Red represents positive correlation and blue represents negative correlation. **(D).** Clustering tree and correlation heat map between different modules. **(E).**Venn diagram of WGCNA-related genes intersecting with DEGs.

### 3.3 Correlation and enrichment analysis of AR-DECSRs

We evaluated the correlation between AR-DECSRs by the Pearson correlation coefficient (Fig. 4A). PPP1R13B was found to be highly correlated with FGFR3 (cor=0.81) and DHCR24 (cor=0.81). DHCR24 was found to be highly correlated with FGFR3 (cor=0.81) and AT4D (cor=0.81). In contrast, CBX8 was found to be lowly correlated with SOD2 (cor=-0.67). Subsequently, we conducted GSVA enrichment analysis for AR-DECSRs, utilizing gene sets (c5.go.v2024.1.Hs.symbol, c2.all.v2024.1.Hs.symbols) from the molecular feature database (MSigDB) as reference. The analysis revealed significant differences between the AR and STA groups across 1917 pathways in the GO dataset (Fig. 4B, Table S6) and 1192 pathways in the KEGG dataset (Fig. 4C, Table S7) (FDR < 0.05, |t value of GSVA score| > 10).

**Fig. 4.**
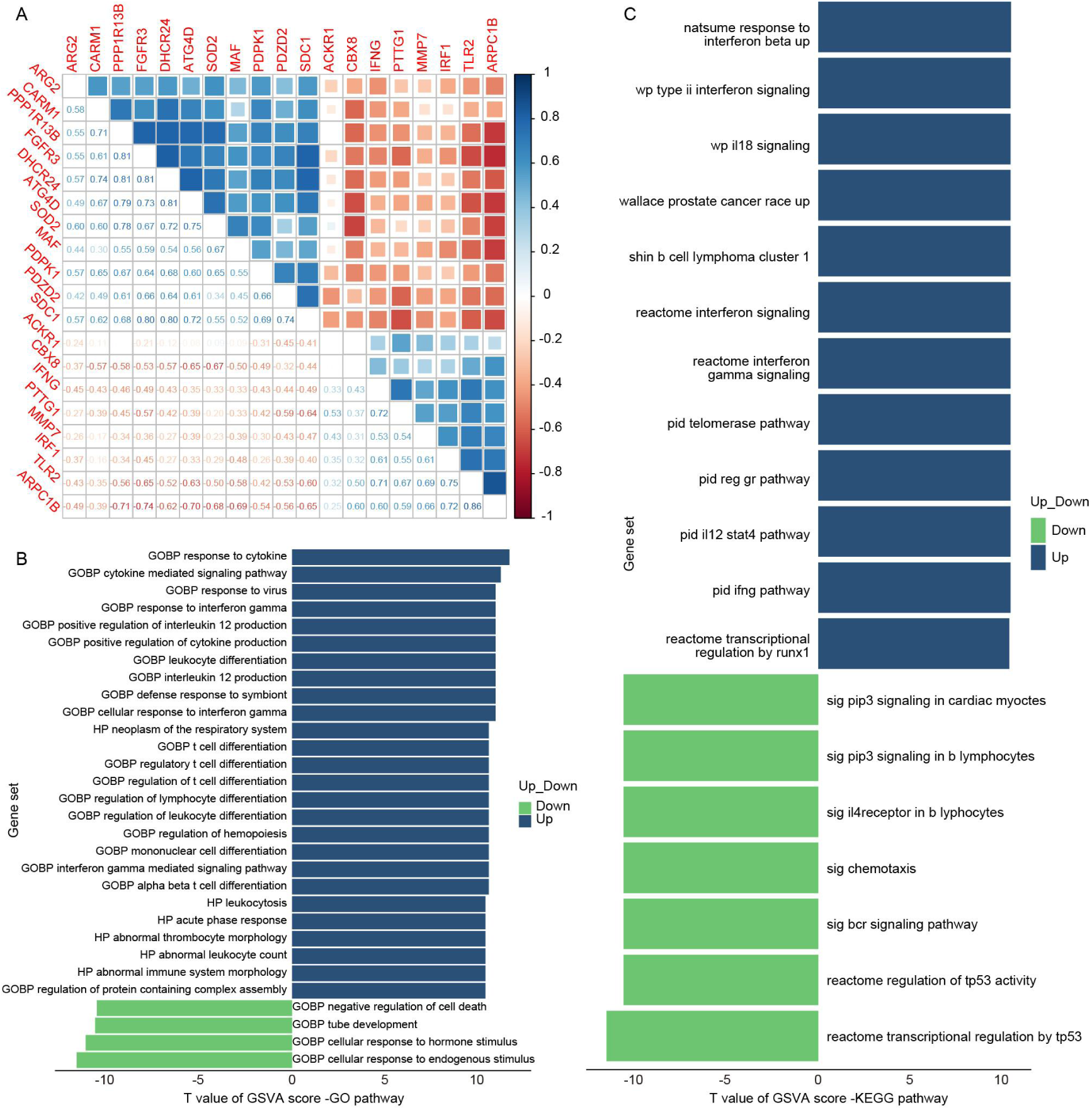
Correlation and enrichment analysis of AR-DECSRs. **(A).** Heatmap of correlation between AR-DECSRs. **(B).** T value of GSVA score of GO-enriched 1917 pathways for GSVA between the AR and ST groups. **(C).** T value of GSVA score of KEGG-enriched 1192 pathways for GSVA between the AR and ST groups.

### 3.4 Identification and validation of hub AR-DECSRs

To improve the accuracy of AR-DECSRs diagnostic AR, we used three machine learning algorithms, Random Forest (Fig. 5A), SVM-RFE (Fig. 5B), and LASSO (Fig. 5C, D), to screen hub AR-DECSRs. We identified a total of four hub AR-DECSRs by combining the results from the three algorithms, including IFNG, CARM1, PDZD2, and ARG2 (Fig. 5E).

**Fig. 5.**
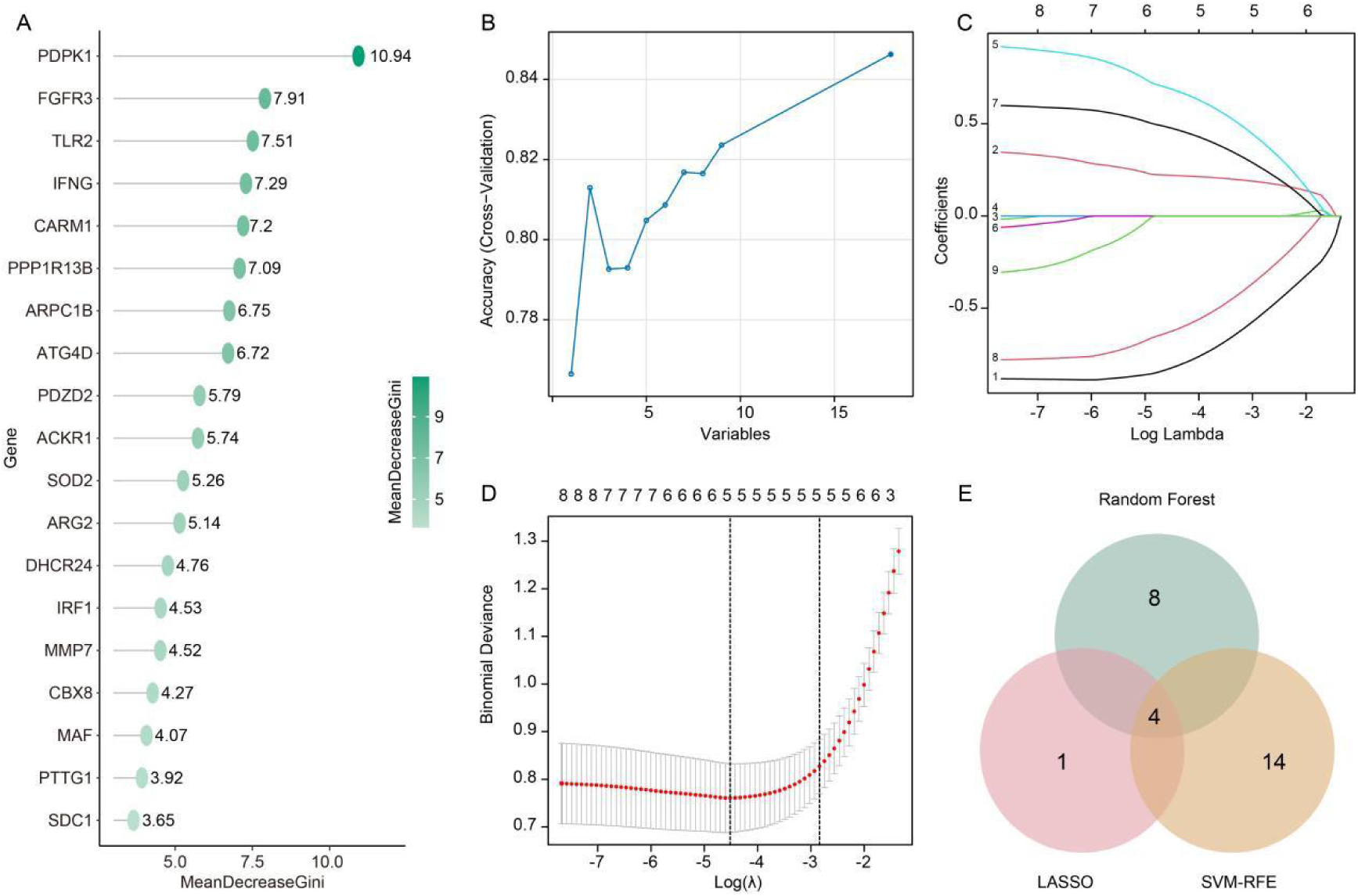
Identification and validation of Hub AR-DECSRs. **(A).** The importance ranking of random forest features. **(B).** Recursive Feature Elimination RFE, the horizontal coordinate represents the number of feature genes and the vertical coordinate represents the generalization error under ten cross-validations. **(C).** Regression coefficient path diagram. X-axis represents the value of the regularisation parameter (Log), while y-axis represents the magnitude of the model parameters. **(D).** LASSO regression cross-validation curves. The x-axis represents the value of the regularisation parameter (log), while the y-axis is the likelihood bias. **(E).** LASSO, Random Forest, and SVM-RFE algorithms for screening Venn diagrams of AR-DECSRs.

### 3.5 Hub AR-DECSRs expression and diagnostic value in AR

Based on the expression levels of hub AR-DECSRs in the GEO-meta dataset and validation sets, we observed that IFNG was significantly upregulated across all AR samples (Fig. 6A-6D). The receiver operating characteristic (ROC) analysis demonstrated that the four Hub AR-DECSRs possess substantial diagnostic value for AR in both the training and validation sets. Among them, IFNG exhibited the highest diagnostic value, AUC=0.81 in the GEO-meta training set (Fig. 6E), AUC=0.79 in the validation set GSE53605 (Fig. 6F), AUC=0.78 in the validation set GSE129166 (Fig. 6G), and AUC=0.71 in the validation set GSE36059 (Fig. 6H). Therefore, IFNG was identified as a candidate gene for subsequent studies.

**Fig. 6.**
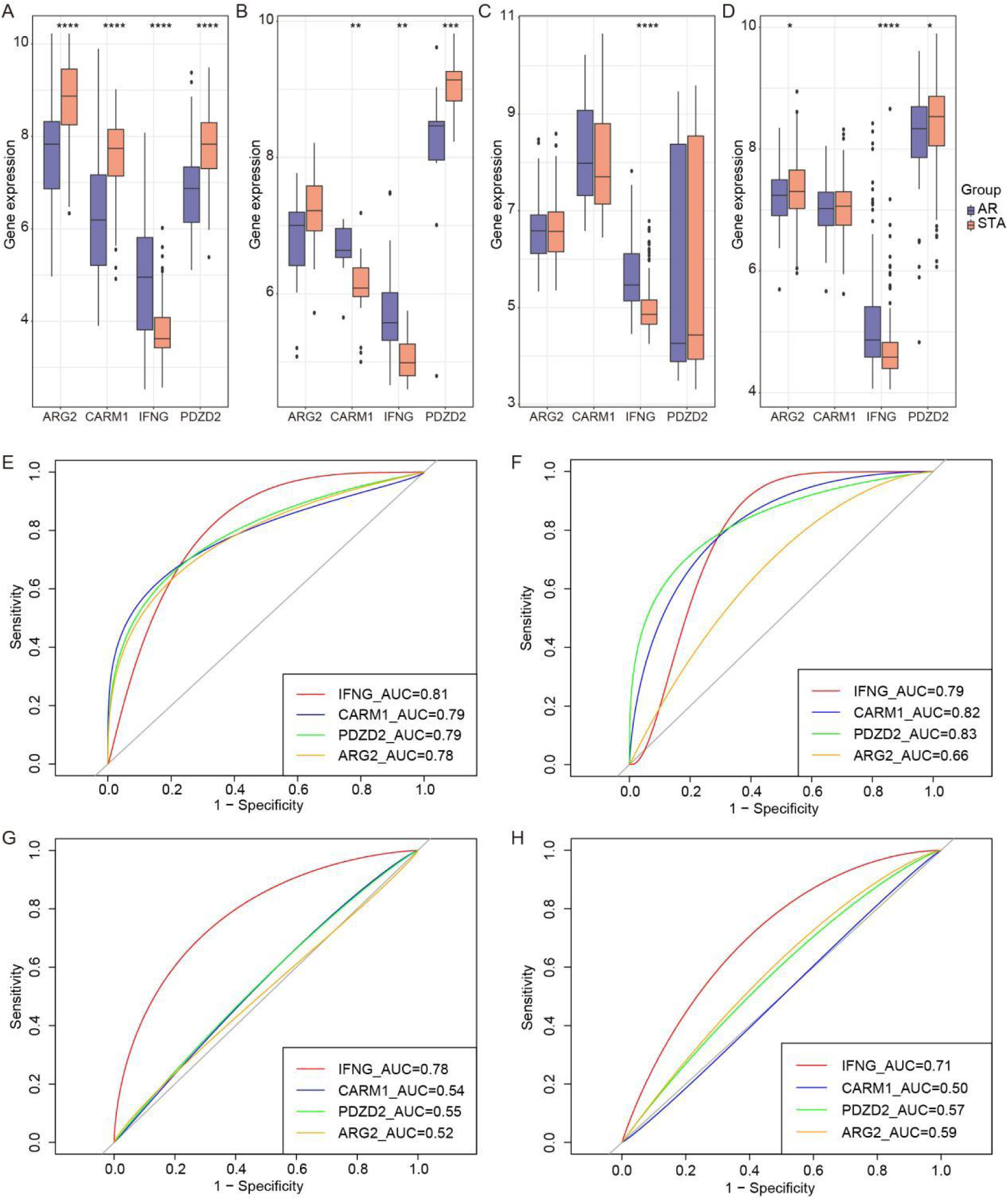
Hub AR-DECSRs expression and diagnostic value. **(A).** The box plot of INFG expression in GEO-meta training set (A),GS53605(B),GSE129166(C), and GSE36059(D). * means *p* < 0.05, ** means *p* < 0.01, *** means *p* < 0.001, **** means *p* < 0.0001.

### 3.6 IFNG was correlated with immune cell infiltration in AR

The infiltration of 28 immune cell types in AR and STA samples was analyzed using ssGSEA. The heatmap depicted an increased enrichment score for each immune cell infiltration in the AR group compared to the STA group (Fig. 7A). Additionally, a correlation analysis among the 28 immune cell types was performed, as illustrated in Fig. 7B. The results revealed significant correlations between most immune cells, e.g. Effector memory CD8 T cell and MDSC were significantly positively correlated (r = 0.92). Subsequently, differential analysis of the 28 immune cell types between AR and STA samples was conducted. Significant differences were observed in all cell types except for Immature Dendritic Cells, Monocytes, and Plasmacytoid Dendritic Cells (Fig. 8A). Furthermore, the differences in 13 immune functions between AR and STA samples were analyzed. The results revealed significant differences in 12 out of the 13 immune functions, with the exception of Type II IFN Response, as shown in Fig. 8B. In addition, we found that IFNG was associated with 26 immune cells (*p*< 0.05, Fig. 8C). IFNG was associated with 12 immune functions between AR and STA samples, such as T.cell.co.inhibition, inflammation.promoting, and check point (*p*< 0.05, Fig. 8D).

**Fig. 7.**
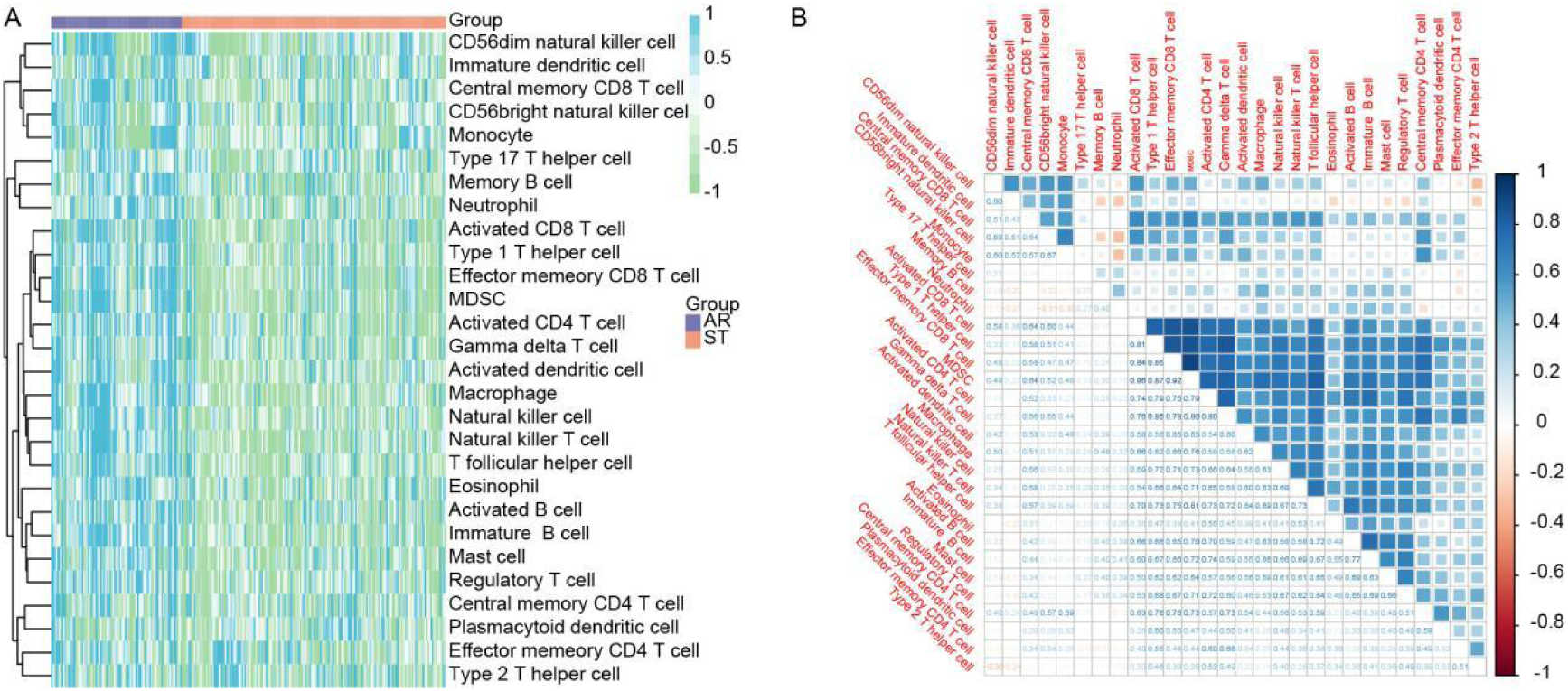
Immune infiltration analysis and correlation analysis among the immune cell. **(A).** Heat map of the enrichment fraction of immune infiltrating cells in AR and STA samples. **(B).** Correlation analysis among 28 immune cells.

**Fig. 8.**
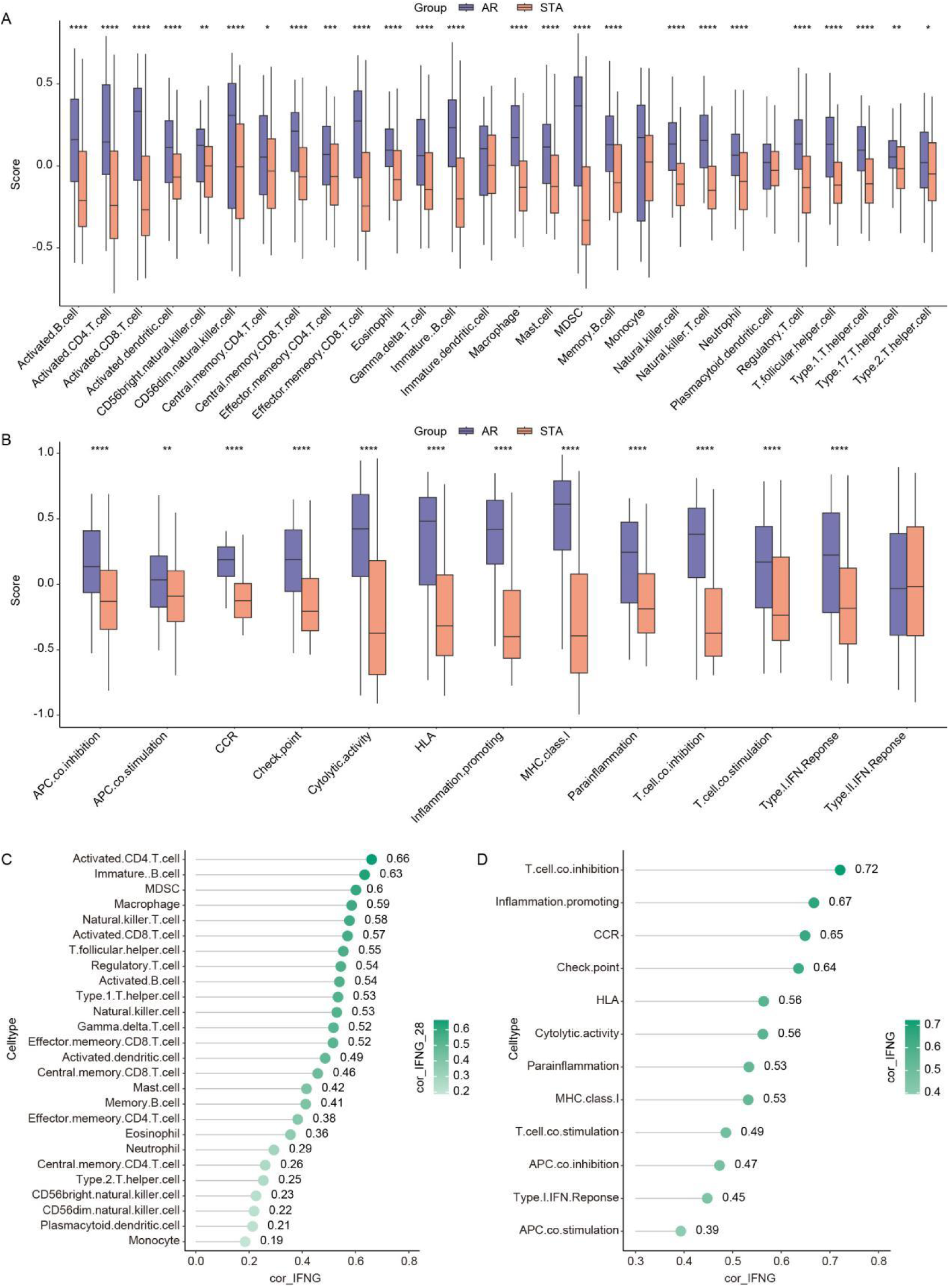
The correlation analysis of immune cell and immune function. **(A)**. Differential analysis of 28 immune cells in AR and STA samples. **(B).** Differences in the function of 13 immune cells in AR and STA samples. **(C)**. Correlation of INFG with 28 immune cells. **(D)**. Fig. 8. The correlation analysis of immune cell and immune function. (A). Differential analysis of 28 immune cells in AR and STA samples. (B). Differences in the function of 13 immune cells in AR and STA samples. (C). Correlation of INFG with 28 immune cells. (D). Correlation of INFG with 13 immune functions. * means p < 0.05, ** means p < 0.01, *** means p < 0.001, **** means p < 0.0001.

### 3.7 Prediction of IFNG-related drugs

We conducted a drug target analysis for the IFNG gene. From the GeneCards database (https://www.genecards.org/), we identified 40 drugs associated with the IFNG gene (Fig. 9A, Table S8). Additionally, using the DGIdb database (version 4.2.0-sha1 afd9f30b, https://dgidb.genome.wustl.edu), we found 28 drugs related to the IFNG gene (Fig. 9B, Table S9), of which 20 drugs have been approved.

**Fig. 9.**
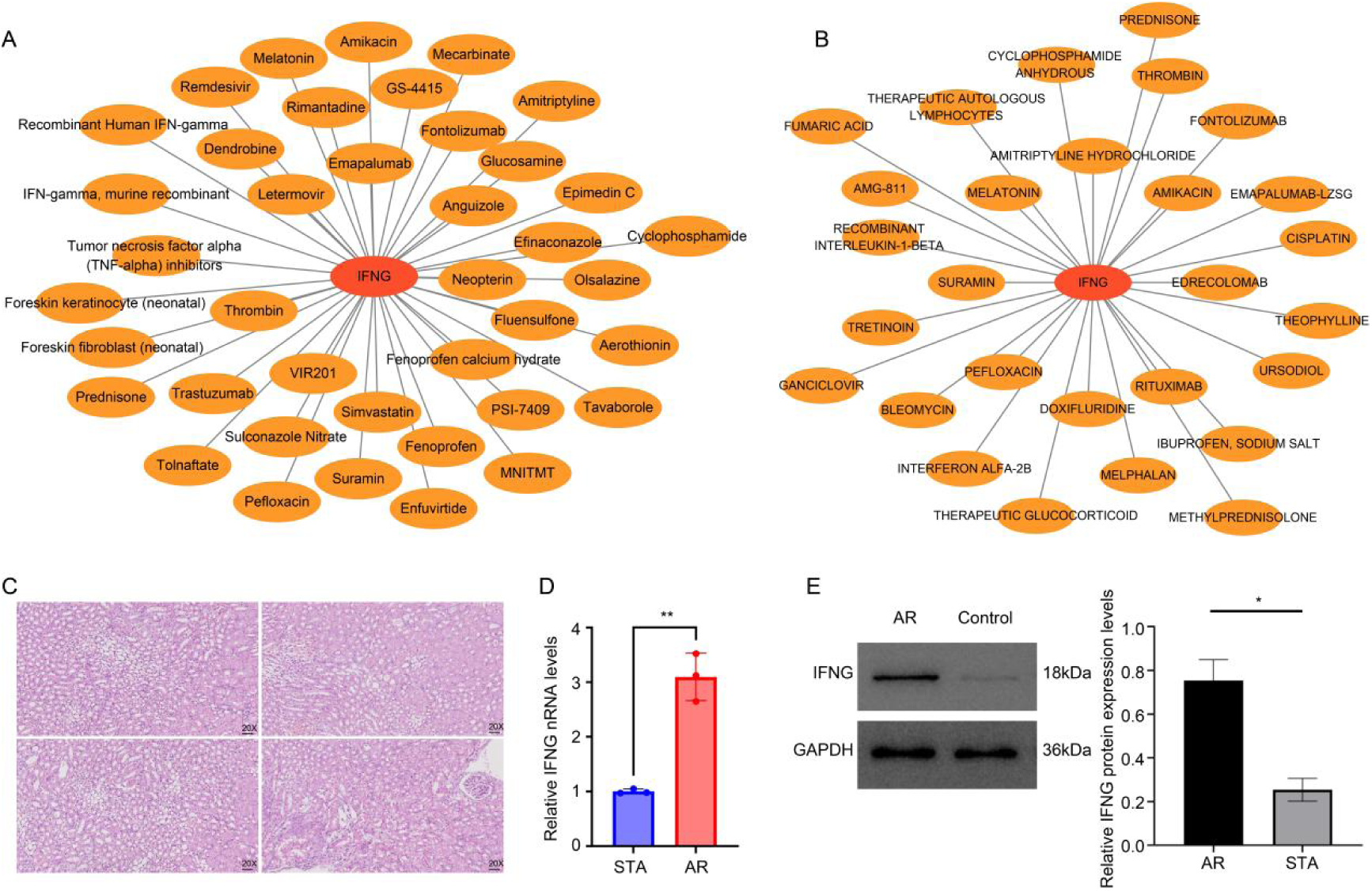
Drug sensitivity and IFNG expression in the kidney tissue of the AR and STA mice. **(A).** Genecards predicts network graphs of drugs and IFNG. **(B).** DGIdb predicts network graphs of drugs and IFNG. **(C)**. Injury of kidney grafts in mice of acute rejection of kidney transplantation. **(D)**. The expression level of IFNG mRNA in the kidney tissue of the AR and STA mice was evaluated by RT-qPCR. **(E)**. The expression level of IFNG protein in the kidney tissue of the AR and STA mice was evaluated by Western blot analysis. ** means *p* < 0.01.

### 3.8 IFNG high expressed in kidney tissues of AR in Allogeneic Kidney Transplant mice

Finally, we established a mouse model of acute rejection in allogeneic kidney transplantation. As illustrated in Fig. 9C, this model exhibited localized lesions in the renal tissue, characterized by abnormal renal tubule morphology, indistinct cytoplasm, and irregular luminal areas, accompanied by minimal infiltration of inflammatory cells. The glomeruli showed signs of dehydration resulting in increased cross-sectional areas. The expression of IFNG mRNA and protein was significantly increased in acute rejection after allogeneic kidney transplantation mice compared to STA mice (Fig. 9D-9E).

## 4 Discussion

Recently, the development of immunosuppressive therapy has significantly increased the incidence of acute allograft rejection in renal transplant patients [28]. However, timely differential diagnosis is still necessary because AR rapidly and progressively impairs graft function [29]. It has been reported that senescence negatively affects organ quality, immunogenicity, recovery from ischemia/reperfusion injury, and transplantation outcomes [30]. In the present study, we employed WGCNA and three machine learning algorithms to identify *IFNG* as the hub cellular senescence-related gene in AR after kidney transplantation. Our findings revealed that *IFNG* was highly expressed in AR samples and could distinguish between STA individuals and AR patients after kidney transplantation. Furthermore, *IFNG* expression was correlated with immune cell infiltration and function, as well as with multiple drugs, suggesting its potential as a biomarker for AR.

Firstly, we discovered that 31 CSRs were differentially expressed between the patients with AR and STA after kidney transplantation. Human studies indicated that the concentration of senescent kidney cells before transplantation is associated with the later development of interstitial fibrosis, tubular atrophy, and chronic allograft nephropathy [31]. The results of GO terms indicated that these 31 genes were involved in cell-substrate junction, focal adhesion, mesonephric epithelium development, and mesonephric tubule development process. These findings indicated that CSRs were correlated with the AR after kidney transplantation.

Moreover, we identified 19 genes correlated with the onset of allergic rhinitis (AR) among 31 CSRs using WGCNA. Through the application of three machine learning algorithms, we identified four genes that were closely associated with the occurrence of AR. Notably, *IFNG* was found to be highly expressed in AR samples and was able to differentiate between STA individuals and AR patients after kidney transplantation. These results were confirmed by previous studies [32, 33].

*IFNG*, also referred to as interferon-gamma (IFN-γ), is a member of the interferon family. It is unique among type II interferons and is distinct from type I interferons, which are primarily produced by plasmacytoid dendritic cells and other immune cells in response to viral infections [34]. *IFN-γ* is produced by various immune cells, including natural killer (NK) cells, natural killer T (NKT) cells, and T lymphocytes, particularly CD4+ Th1 cells and CD8+ cytotoxic T cells [35]. As a pleiotropic cytokine, *IFN-γ* exhibits antitumor [36], antiviral [37], and immunomodulatory functions [38]. It has been reported that molecules expressed in effector T cells, antigen-presenting cells (macrophages, dendritic cells, B cells), and *IFN-γ* induced genes predominated in TCMR landscape in kidney transplant [39].

In the present study, *IFN-γ* expression was correlated with Treg, macrophage, CD4+ T cells, and CD8+ T cells in AR after kidney transplantation. *IFN-γ* MSCs have been demonstrated to inhibit the infiltration of inflammatory cells, including T cells and macrophages, by inducing Treg cells. This leads to pronounced anti-fibrotic effects in renal ischemia reperfusion injury (IRI) models [40]. In kidney transplant recipients, DNA methylation of *IFN-γ* in memory CD8+ T cells increased within the first three months after transplantation [41]. Moreover, *IFN-γ* could promote the polarization of macrophages to the M1 phenotype, which was associated with pro-inflammatory activities and can contribute to tissue damage in the allograft [42]. In ABMR, M1 macrophages had the highest *IFN-γ* receptor levels and were heavily infiltrated in the allografts [42]. Collectively, *IFN-γ* was closely associated with the activation of various immune cells and the enhancement of inflammatory responses in the AR of kidney transplants and. Monitoring the production of *IFN-γ*, along with changes in related immune cell populations, might provide critical insights for the diagnosis and treatment of renal transplant rejection.

## 5. Conclusion

The present study identified cellular senescence-associated gene *IFNG* plays a significant role in AR following kidney transplantation. Our findings demonstrate that IFNG is highly expressed in samples from patients experiencing AR and serves as a robust biomarker that can effectively differentiate between individuals with stable kidney function and those with AR. Additionally, the expression levels of *IFNG* were found to be intimately linked with the infiltration and functionality of immune cells, suggesting its potential involvement in the immunological processes underlying allograft rejection. The correlation of *IFNG* with various drugs implies that it may be a target for therapeutic intervention to mitigate AR episodes. Collectively, our results highlight *IFNG* as a promising biomarker and potential therapeutic target for the management of acute rejection in kidney transplant recipients

## Declarations

## Acknowledgement

Not applicable.

## Ethical approval and consent to participate

The conduct of all animal-related experiments adhered to the National Guidelines for Ethics Review of Laboratory Animal Welfare. Ethical approval for all mouse experiments was granted by the Laboratory Animal Ethics Committee of Beijing MDKN Biotech Company (MDKN-2023-054).

## Consent for publication

Not applicable.

## Data Availability Statement

Data supporting the conclusion of this study are openly available in the Gene Expression Omnibus (GEO, https://www.ncbi.nlm.nih.gov/geo/) database.

## Authors’ contributions

Changhai Xu participated in the design of this study, and Qin Feng performed statistical analysis. Xueying Wang, Haibo Wu, Wei Li, Fei Lin, Na Lin, Shiyin Shen, Shubin Pan, Tong Chen, Donghui Zhang and Long He carried out the study and collected background information. Yan Cui drafted this manuscript. All the authors have read and approved the final manuscript.

